# Neurite outgrowth deficits caused by rare PLXNB1 mutation in pediatric bipolar disorder

**DOI:** 10.1101/2022.05.06.22274499

**Authors:** Guang Yang, Ethan Parker, Bushra Gorsi, Mark Liebowitz, Colin Maguire, Jace B. King, Hilary Coon, Melissa Lopez-Larson, Jeffrey Anderson, Mark Yandell, Alex Shcheglovitov

## Abstract

Pediatric Bipolar Disorder (PBD) is a severe mood dysregulation condition that affects 0.5–1% of children and teens in the United States. It is associated with recurrent episodes of psychosis and depression and an increased risk of suicidality. However, the genetics and neuropathology of PBD are largely unknown. Here, we used a combinatorial family-based approach to characterize the cellular, molecular, genetic, and network-level deficits associated with PBD. We recruited a PBD patient and three unaffected family members from a family with a history of psychiatric illnesses. Using resting-state functional magnetic resonance imaging (rs-fMRI), we detected altered resting-state functional connectivity in the patient as compared to the unaffected sibling. Using transcriptomic profiling of patient and control induced pluripotent stem cell (iPSC) derived telencephalic organoids, we found aberrant signaling in the molecular pathways related to neurite outgrowth. We corroborated the presence of neurite outgrowth deficits in patient iPSC-derived cortical neurons and identified a rare homozygous loss-of-function *PLXNB1* variant (c.1360C>C; p.Ser454Arg) in the patient. Expression of wild-type *PLXNB1*, but not the variant, rescued neurite outgrowth deficit in patient neurons, and expression of the variant caused neurite outgrowth deficit in cortical neurons from *PlxnB1* knock-out mice. These results indicate that dysregulated PLXNB1 signaling may contribute to an increased risk of PBD and other mood dysregulation-related disorders by disrupting neurite outgrowth and functional brain connectivity. Overall, the study established and validated a novel family-based combinatorial approach for studying cellular and molecular deficits in psychiatric disorders.

## Introduction

Bipolar disorder (BD) is a severe psychiatric illness that affects ∼1-2% of the world population(1,2). The hallmark symptoms of bipolar disorder are episodes of clinical mania and depression. However, many symptoms that patients experience are nonspecific and overlap with other psychiatric illnesses, including schizophrenia, major depressive disorder, and attention deficit hyperactivity disorder (3,4). Concordantly, there exists a significant overlap in the genetic risk factors that underlie bipolar disorder and other psychiatric illnesses, such as major depressive disorder and schizophrenia (5–7). Pediatric bipolar disorder (PBD) is an early on-set BD diagnosed before age 18. It is estimated to affect 0.5–1% of children and teens in the United States but is highly underdiagnosed due to substantial overlap with the diagnoses of other mood dysregulation disorders (8,9). PBD has been associated with increased risks for suicidal behavior, substance abuse, problems with authority, and hospitalizations.

Despite the tremendous socioeconomic burden of BD and PBD, the underlying pathology and genetics of these disorders remain largely unknown. This is partly due to the lack of animal models that recapitulate the complex symptom profile of patients (10). Neuroimaging studies in patients with BD and PBD have identified abnormalities in cortical and subcortical neural circuits involved with emotional processing and cognitive control (11–13); however, it is unknown how the detected circuit-level abnormalities may reflect underlying changes in the brain at the cellular and molecular levels. Induced pluripotent stem cell (iPSC)-derived neurons from BD patients and healthy individuals have been used to study the cellular and molecular deficits that are disrupted in association with BD (14). It has been demonstrated that BD is associated with reduced proliferation of neural progenitor cells, impaired neuronal differentiation (15,16), impaired cell adhesion (17), and neuronal hyperexcitability (18,19), which could be modulated by lithium responsiveness (17–19). However, despite the progress in uncovering the cellular deficits associated with BD, the genetic underpinnings of the cellular deficits have not been determined, and it is unknown whether patients recruited for iPSC-based studies also show abnormalities in neural circuitry.

BD is one of the most heritable psychiatric disorders (20), with an estimated heritability of 60-85% (21,22). Genome-wide association studies (GWAS) have been used to identify common variants associated with BD (23–27). These studies have uncovered hundreds to thousands of common low-risk variants that may each contribute a small amount of risk for bipolar disorder, including variants in genes encoding calcium and potassium channels, neurotransmitters, cell-adhesion proteins, and other proteins that regulate neuronal functions and excitability. This information has been used to examine the developmental stages associated with these disorders (28), and efforts are underway to integrate GWAS variants with expression quantitative trait loci data to identify candidate causal variants for neuropsychiatric disorders (29). However, the impacts of GWAS variants on the cellular and molecular properties are difficult to study due to small effect sizes that are unlikely to yield measurable cellular and molecular phenotypes.

In addition to common variants, damaging rare and de novo variants have also been identified in association with BD (25,26,30,31); Unlike common variants of low effect sizes, rare variants conferring relatively larger genetic risk are particularly suitable for investigation with iPSC-based models as they can be pinpointed to specific genes and are amenable to genetic modeling (21). Indeed, rare, high-impact gene-coding and structural variants have been discovered to contribute causally to brain-related pathology in patients with psychiatric illnesses (32–34) and may also contribute to the severity of psychiatric illness (35). Understanding how damaging rare or de novo variants could contribute to cellular phenotypes in iPSC-derived neurons may provide a window into the pathophysiology of bipolar disorder and other major psychiatric disorders.

In the present study, we have developed a novel approach to identifying and functionally interrogating rare variants in patients with severe psychiatric disorders. We recruited and characterized a family that included an individual with PBD. We performed rs-fMRI and identified functional connectivity differences between the patient and the sex-matched unaffected sibling consistent with the results of our previous study on a larger cohort of individuals with PBD (36). We generated iPSCs and iPSC-derived neurons and organoids from all family members and found that patient iPSC-derived neurons exhibited reduced neurites as compared to control neurons. Finally, we identified a rare loss-of-function homozygous mutation in *PLXNB1* in patient cells that was responsible for the neurite outgrowth deficits. Collectively, these results suggest that loss-of-function mutations in *PLXNB1*, which lead to impaired neurite outgrowth, can contribute to disrupted resting-state functional connectivity and PBD risk.

## Methods

### Study Participants

The Institutional Review Board at the University of Utah approved this study. The family participating in this study was recruited via clinical referral. All subjects provided written informed consent prior to study participation. The patient and the unaffected sibling underwent clinical and diagnostic interviews by a board-certified psychiatrist (MLL) using the Mini-international Neuropsychiatric Interview (37). The patient received a diagnosis of bipolar disorder at age 12. The Child Behavioral Checklist (CBCL) (38), which is a parent-report assessment that measures behavioral and emotional problems related to psychopathology in children, was completed by the participant’s mother. The full-scale intelligence quotient was assessed using the Wechsler Abbreviated Scale of Intelligence (39).

### Functional magnetic resonance imaging

Images were acquired on a Siemens 3T Prisma scanner using a 64-channel head coil. The scanning protocol consisted of an MP2RAGE acquisition for an anatomic template (TR=5000 ms, TE=2.93 ms, TI_1_=700 ms, TI_2_=2030 ms, flip angle_1_=4°, flip angle_2_=5°, field of view=256 mm, 176 slices, resolution=1 × 1 × 1 mm, sagittal acquisition) and three ∼15 minute resting state acquisitions using multiband BOLD echo-planar images (each scan contained 1220 volumes, TR = 730 s, TE = 33.2 ms, multiband factor=8, 2×2×2 mm resolution). During resting state scanning, subjects were instructed to “Keep your eyes open and remain awake and try to let thoughts pass through your mind without focusing on any particular mental activity.”

Despiking (40) was performed using AFNI software package (41) (3dDespike) for initial correction of head motion displacement. Motion correction (realign), coregistration to MP2RAGE, segmentation of MP2RAGE, and normalization of MP2RAGE and BOLD to MNI template was performed in SPM12b (Wellcome Trust, London) for MATLAB (Mathworks, Natick MA). Phase-shifted soft tissue correction (PSTCor) (42) was used to regress physiological waveforms as well as regressors obtained from subject motion parameters, degraded white matter, degraded CSF, and soft tissues of the face and calvarium. No regression of the global signal was performed to avoid contamination of gray matter sources (42–44). Censoring of frames showing greater than 0.2 mm (motion scrubbing) was performed as a final step prior to analysis with concatenation of remaining frames (45).

Differences in connectivity were obtained for the largest 25 clusters from the Yeo et al. brain parcellation as previously described (46,47) with connectivity difference represented by the functional connectivity of the bipolar sibling – control sibling. Differences in functional connectivity between the siblings are displayed below for each pair of 25 cortical brain regions.

The histogram of functional connectivity in the pediatric bipolar and healthy sibling in Fig 1D was generated by parcellating the brain to 6923×6923 regions of interest covering cortical and subcortical gray matter. Functional connectivity values for each unique pair of connections was generated (48,49) and plotted.

**Figure 1.**
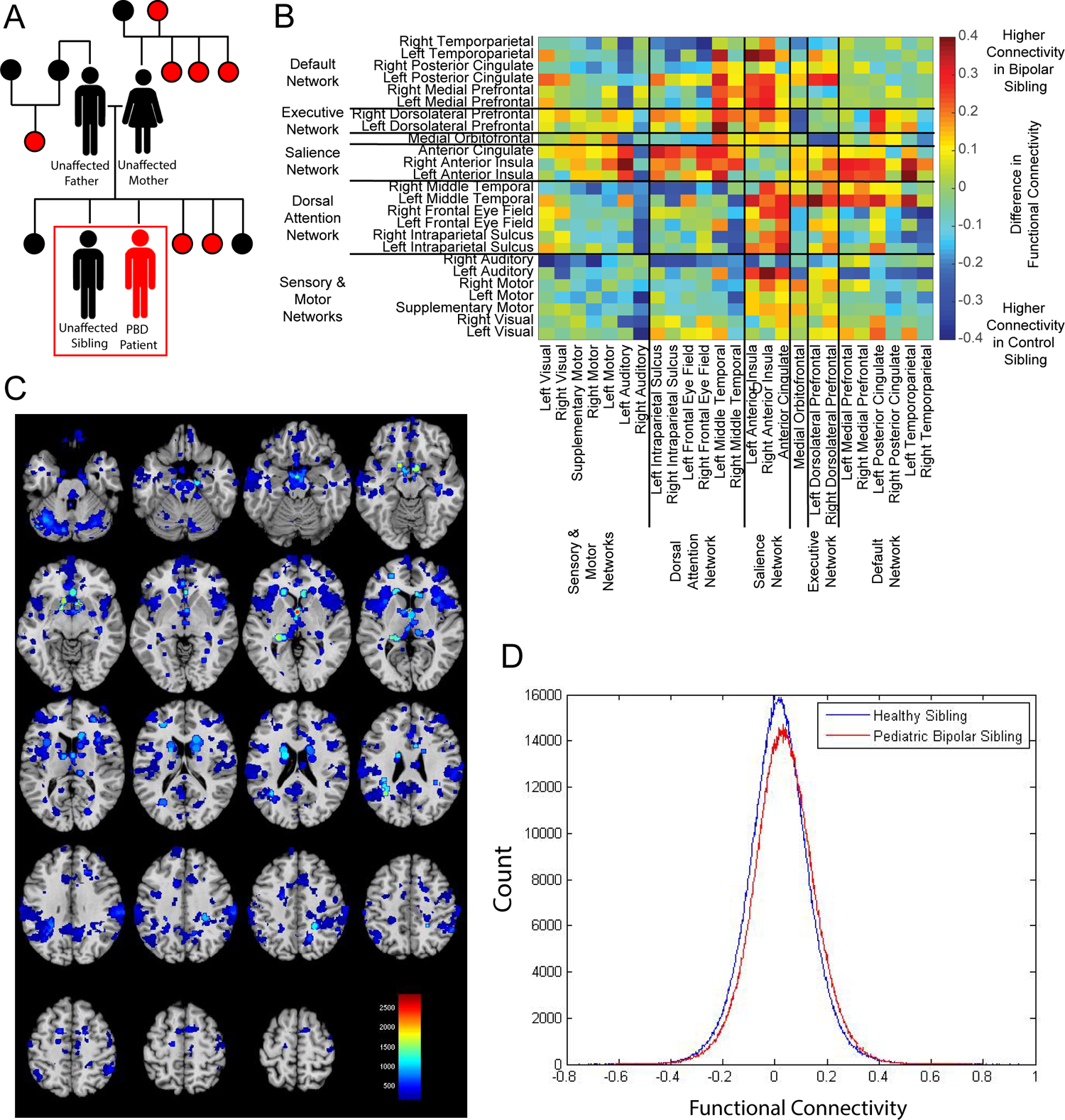
Functional brain connectivity differences in an individual with PBD. A. Schematic of quartet recruited into the present study and extended pedigree. A family quartet, consisting of patient diagnosed with PBD and unaffected mother, father, and sibling, were recruited for this study. Sex and sibling orders in the extended family are not disclosed. Solid red circles indicate relatives with a history of psychiatric symptoms or diagnosis, as reported through parental interviews. Patient and unaffected sibling of the same sex and similar age were subjected to rs-fMRI. B. Heatmap matrix depicting differences between unaffected sibling and patient in functional connectivity between 25 different cortical brain regions organized into 5 networks. C. Heatmap depicting connectivity difference in different brain regions between unaffected sibling and patient. D. Histogram of functional connectivity values in between each unique connection of 6923 parcellated regions of interest for patient and unaffected sibling.

### Whole Exome Sequencing

DNA samples were extracted using the Perkinelmer Chemagic 360 DNA extraction kit and subjected to whole exome sequencing (WES). The four samples with Kindred ID BP-Fam01 were sequenced at University of Utah HCI Core and 125 Cycle Paired-End WES was performed on the Illumina HiSeq 2000 (University of Utah).

### Alignment and Variant Calling

Alignment and variant calling were performed by the Utah Center for Genetic Discovery (UCGD) core services. Fastq files from paired-end sequencing were aligned to the human reference build GRCh38 using BWA-MEM (Burrows-Wheeler Aligner). Variants were called through the UCGD pipeline using the Sentieon software package (https://www.sentieon.com) (50). SAMBLASTER was used to mark duplicate reads and de-duplicate aligned BAM files. The following algorithms from the Sentieon software package3 Realigner and Qual Cal were utilized for INDEL realignment, base recalibration to produce polished BAM files. Polished BAM file was processed using the Sentieon’s Haplotyper algorithm to produce gVCF files (51). Sample gVCF files were combined and jointly genotyped with 287 unrelated Utah samples from the 1000 Genomes Project (CEU) samples to produce a multi-sample VCF file.

### Quality Control

We applied several quality control algorithms to the Fastq files, BAM files and VCF files (Ewels 2016). Read quality, read duplication rate, presence of adapter and overrepresented sequences in Fastq files were extracted from the Fastp files (52). We estimated depth and coverage of aligned sequence data from BAM indexes using IndexCov (53). Sample qualities in final VCF were evaluated using Peddy (54) to determine and confirm sex, relatedness, heterozygosity of each individual to identify any potential sample quality issues.

### Identification of damaging variants

A Pedigree Variant Annotation, Analysis, and Search Tool (pVAAST), version 2.2.0, (55) was performed on four individual samples (cases) and 2492 background controls from the 1000 Genomes Project to test for association between genetic burden and phenotype. pVAAST scores variants based on their functional variant prediction and evidence of co-segregation among affected family members. Analysis excluded low-quality variants with statistically significant departure from Hardy-Weinberg equilibrium (*P* < .0001) or with missing genotype rates greater than 10.0% among background genomes. Genes were then ranked according to an empirical p-value determined by running a permutation test against a set of background control samples and counting the number of controls that have a genotype at least as damaging as that observed in the affected family members. pVAAST was run with 1×10^4^ permutations against the 2492 background genomes from the 1000 Genomes Project. Additional re-ranking of pVAAST outputs occurred using Phenotype Driven Variant Ontological Re-ranking tool (PHEVOR) (56) which uses Human Phenotype Ontology (HPO) terms and connections to Gene Ontology (GO) terms to re-prioritize potentially damaging alleles. We used the following HPO terms: Bipolar affective disorder HP:0007302, Behavioral abnormality HP:0000708, Abnormal emotion/affect behavior HP:0100851.

### Bulk RNA Sequencing and Analysis

Total RNA was purified using the RNeasy Micro Kit (Qiagen) with DNase I digestion. RNA was purified from three organoids from each family member (Patient, unaffected sibling, mother, father). Libraries were prepared using the NEBNext Ultra II Directional RNA Library Prep Kit with rRNA depletion and verified using the Bioanalyzer RNA Nano Assay (Agilent) before sequencing on the NovaSeq 6000 (Illumina) with paired-end 150b reads. Optical duplicates were removed with Clumpify v38.34. Adapter sequences were trimmed with Cutadapt and reads were aligned to the reference genome (GRCh38) using STAR version 2.7.3a. Features were counted using FeatureCounts version 1.6.3. DESeq2 was used to perform differential expression analysis. Gene set enrichment analysis (GSEA) was performed using the GSEA 4.1.0 GSEAPreranked tool (57) with default parameters against the following Gene Ontology dataset domains: GO Biological Processes, GO Molecular Functions, GO Cellular Component. Ranked gene lists used for the GSEA analysis was generated using the write_gsea_rnk function from the hciR R package (https://github.com/HuntsmanCancerInstitute/hciR) for the following contrasts: patient vs. unaffected sibling, patient vs. mother, patient vs. father, and patient vs. all controls pooled.

For the BrainSpan transcriptome correlational analysis, we combined normalized gene expression data from all patient and control SNR-derived organoids with BrainSpan dataset (http://www.brainspan.org) consisting of 309 samples from 27 different donors ranging from 8pcw to 1 year. Spearman’s correlation coefficient was calculated between each organoid sample and each BrainSpan sample for dorsolateral prefrontal cortex, anterior (rostral) cingulate (medial prefrontal) cortex, and ventrolateral prefrontal cortex average gene expression.

Bulk RNA sequencing deconvolution analysis was performed using the Cell Population Mapping algorithm (58) with default parameters. Relative abundance of neural progenitor cells, excitatory neurons, and inhibitory neurons were inferred from Bulk RNA sequencing data from all patient and control 1-month old SNR-derived organoids using single-cell RNA sequencing data from 1-month old SNR-derived organoids (59).

### Immunostaining

For all immunostaining experiments, cells were first washed with 1x PBS then fixed with 4% paraformaldehyde for 15 minutes at room temperature. Cells were then washed 3x with PBS-Glycine. Then, cells were permeabilized with 0.3% Triton X-100 and blocked for 1 hour with 3% BSA. Cells were then incubated with primary antibody diluted in 3% BSA for either 1 hour at room temperature or overnight at 4C. Cells were then washed 3 times with PBS, then incubated with secondary antibodies. Coverslips were then mounted on glass slides using VectaShield (Vectorlabs). The following antibodies were used: Ms α TuJ1 (1:1000, BioLegend/Covance MMS-435P), Ch α MAP2 (1:1000, Novus Biologicals NB300-213), Ch α GFP (1:1000, Novus Biologicals NB100-1614), Ms α PlexinB1 (1:500 Santa Cruz Biotechnology sc-28372), Rat α Ki67 (1:1000, Thermo Scientific 14-5698-82), Rb α Tbr1 (1:1000, Abcam ab31940), Ms α Satb2 (1:1000, Abcam ab51502), Rat α Ctip2 (1:1000, Abcam ab18465), Ms α GAD67 (1:1000 EMD Millipore MAB5406), Phalloidin-488 (1:1000, AAT Bioquest 23153).

### iPSC Generation and Characterization

Blood samples were obtained from all four family members (father, mother, unaffected sibling, patient). Induced pluripotent stem cells (iPSCs) were generated from patient and control fibroblasts using the CytoTune-iPS 2.0 Sendai Reprogramming Kit. Characterization of iPSCs were performed using immunohistochemistry. iPSCs were stained and assessed for pluripotency markers Oct4, Sox2, Nanog, and Tra-1-60. (Supplementary Fig 1). Karyotyping of iPSCs from all family members was performed using the KaryoStat Assay (ThermoFisher) to assess for the presence of copy number variants (Supplementary Fig 2).

### iPSC Culture and Neural Differentiation

Human pluripotent stem cell lines used in this study were maintained on Matrigel (1%, BD) in 1:1 ratio of Essential 8 Medium (Thermo Fisher Scientific) and StemFlex Medium (Thermo Fisher Scientific). For neural differentiation, cells were passaged at high density to reach ∼90% confluency 2-3 days post-passaging. For induction, E8 medium was replaced with neural differentiation (ND) medium containing a 1:1 mixture of N2 medium (DMEM/F-12, 1% N2 Supplement [Cat#17502048], 1% MEM-NEAA, 2 µg/ml Heparin, and 1% Pen/Strep) and B-27 medium (Neurobasal-A [Cat#10888022], 2% B27 Supplement with vitamin A [Cat#17504044], 1% GlutaMAX and 1% Pen/Strep) supplemented with SMAD inhibitors (4 µM dorsomorphin and 10 µM SB431542). ND medium was replaced daily for the next 7–10 days. On days 7-10, cells were dissociated with dispase. Cell clumps were re-plated on Matrigel-coated plates in ND medium with EGF (10 ng/mL) and FGF (10 ng/mL). Approximately 50–100% of media was replenished daily for the next 3–6 days to remove dying cells. Neural rosette clusters appeared after ∼6 days in EGF/FGF-containing media. Neural cells are expanded for an additional 1-2 weeks then dissociated into single cell suspension with Accutase (STEMCELL Technologies 07920) and plated on Matrigel-coated plates in 1:1 mixture of N-2 and B-27 medium. For experiments involving iPSC-derived neurons, neurons were dissociated with papain (20U/mL, Worthington) and plated on coverslips coated with Poly-L-Ornithine (100ug/mL, Sigma-Aldrich P3655-100MG) and Laminin (50ug/mL, Sigma-Aldrich L2020-1MG)

### Generation of Organoids

For generation of organoids used in bulk RNA sequencing, iPSCs were first cultured and induced into generate neural rosettes. Single neural rosettes (SNRs) were mechanically isolated from plates of neural rosettes using a sterile glass hook and re-plated on Matrigel-coated plates at low density. On days 15–19, SNRs were manually isolated and transferred to ultra-low attachment plates in ND medium with EGF (10 ng/mL) and FGF (10 ng/mL). Plates were kept on an orbital shaker (45 rpm) in a 37°C, 5% CO_2_ incubator with 50% media exchange every other day to maintain the final concentrations of EGF and FGF at 10 ng/mL. Brightfield images of organoids were taken every 7 days. On day 30 post-induction, final images of organoids were taken and organoids that are visually similar in size and appearance were harvested for bulk RNA sequencing.

### Neurite Outgrowth Assay

Patient and control iPSC-derived neurons (1-month post-induction) were dissociated with papain and plated on 15mm coverslips coated with Poly-L-Ornithine (100ug/mL, Sigma-Aldrich P3655-100MG) and Laminin (50ug/mL, Sigma-Aldrich L2020-1MG) at a density of 70,000 cells/coverslip. Neurons were incubated in neuronal media (1:1 mixture of N-2 and B-27) for 48 hours. Cells were then fixed with 4% paraformaldehyde and stained for DAPI and TuJ1 for neurite length quantification. Images were obtained at 20x magnification using either the EVOS M5000 microscope or the Nikon A1R confocal microscope. Semi-automated neurite length quantification was performed using HCA-Vision (CSIRO).

### Generation of PLXNB1 Plasmids

A PCDNA3.1 vector containing full-length human PLXNB1 was obtained from Dr. Xuewu Zhang. T2A-EGFP was cloned into the PCDNA3.1-PLXNB1 plasmid to generate wild-type PLXNB1-T2A-EGFP plasmid. Patient-specific PLXNB1 1360 A>C point mutation was generated from the wild-type PLXNB1-T2A-EGFP plasmid using QuikChange Mult Site-Directed Mutagenesis Kit (Agilent Technologies). Confirmation of the point mutation was performed using PCR and Sanger Sequencing.

### Growth Cone Collapse Assay

iPSC-derived neurons (1-month post-induction) were dissociated with papain and plated on chamber slides coated with Poly-L-Ornithine and Laminin. 48 hours post-plating, neurons were treated with either vehicle control (PBS) or 8nM Sema4D for 1 hour. Cells were then fixed with 4% paraformaldehyde and stained with TuJ1, Phalloidin-488, and DAPI for growth cone evaluation. The percentage of neurites containing growth cones was quantified using ImageJ. All analyses were performed blind.

### Functional Characterization of PLXNB1 Ser454Arg

The Cos7 collapse assay was used to functionally characterize PLXNB1 Ser454Arg. COS7 cells were cultured in DMEM/F12 (Invitrogen) supplemented with 10% FBS and 1% Pen/Strep. On day 0, cells were replated onto tissue culture-treated 24-well plates at a concentration of 15,000 cells/well. On day 1, cells were co-transfected with 0.5ug mKate and 1ug of either wild-type PLXNB1-T2A-EGFP or PLXNB1-Ser454Arg-T2A-EGFP using Lipofectamine 3000 Transfection Reagent (Thermo Fisher Scientific L3000008). On day 3, cells were treated with 2nM Sema4D for 30 minutes, then fixed with 4% paraformaldehyde. Images of cell morphology were taken at 40x magnification using the EVOS M5000 microscope. Cells were categorized as “collapsed” or “non-collapsed” based on morphology of RFP+ signal.

### Neurite Outgrowth Rescue in iPSC-derived neurons

iPSC-derived neurons (14 days post-induction) from patient and unaffected sibling were dissociated with Accutase (STEMCELL Technologies 07920) and resuspended in neuronal medium (1:1 mixture of complete N-2 and B-27 medium. Dissociated cells were electroporated with either 5ug of wild-type PLXNB1-T2A-EGFP or PLXNB1-Ser454Arg-T2A-EGFP using the ECM 830 Square Wave Electroporation System (BTX) (340V, 900us, 1 pulse) and let rest at room temperature for 10 minutes. Cells were then transferred to poly-L-ornithine and laminin-coated coverslips at a density of 200,000 cells/well. 12 hours later, half the media was replaced with 1:1 mixture of N-2 and B-27 medium supplemented with 10nM BDNF(Novus Biologicals NBP1-99674-50ug) and 1% Pen/Strep. 48 hours later, cells were fixed with 4% paraformaldehyde and immunostained with antibodies against GFP and TuJ1. Images of neurons were taken at 20x magnification using the EVOS M5000 microscope. Neurite length quantifications were assessed using the ImageJ plugin NeuronJ (60).

### Mouse experiments

PlexinB1 +/- mice were generated as described by Friedel et al. 2005 and were received as a gift the Paradis lab at Brandeis University. Mating pairs between PlexinB1 +/- mice were set up to produce PlexinB1 -/- mice for all neuronal culture experiments. Cortical neurons were harvested at P0 and dissociated using papain (Worthington). Neurons were electroporated with the ECM 830 Square Wave Electroporation System (BTX) with either 5ug of wild-type PLXNB1-T2A-EGFP or mutant PLXNB1-T2A-EGFP plasmid, then plated onto poly-L-ornithine and laminin-coated 10mm coverslips at a density of 250,000 cells/coverslip. Cells were cultured in MEM (Gibco) supplemented with 10% FBS, 1M Glucose, 200mM GlutaMax (gibco), and 100mM pyruvic acid (sigma). 12 hours later, media was changed to Neurobasal A (Gibco) supplemented with B27, 1% Pen/Strep, 200mM GlutaMax, and 1M Glucose. 48 hours later, cells were fixed and immunostained. 48 hours later, cells were fixed with 4% paraformaldehyde and immunostained with antibodies against GFP and TuJ1. Images of neurons were taken at 20x magnification using the EVOS M5000 microscope. Neurite length quantifications were assessed using the ImageJ plugin NeuronJ (60).

## Results

We ascertained an individual with PBD in a Utah family with multiple individuals affected by common psychiatric illnesses, including bipolar disorder, anxiety, and depression (Fig 1A), suggesting a potential contribution of genetic factors to the psychiatric disorders in this family. To study the biological basis of PBD, we recruited a quartet from this family consisting of an unaffected father, unaffected mother, unaffected male sibling, and male diagnosed with PBD at age 12. The patient and unaffected sibling were both adolescents at the time of recruitment. Participants under the age of 18 underwent a clinical and diagnostic semi-structured interview by a board-certified child psychiatrist (MLL). The Child Behavioral Checklist (38) was assessed for the patient and unaffected sibling (Supplementary Table 1). The patient scored within the clinical range for the category “CBCL Internalizing Problems”. Both individuals scored within the normal range for “CBCL Externalizing Problems” and displayed normal IQ (Supplementary Table 1) (39).

We previously demonstrated that PBD is associated with abnormal functional connectivity between the default mode network (DMN) and salience network (SN) using rs-fMRI (36). To investigate the differences in the resting-state functional connectivity between the patient and unaffected sibling, we performed multiple high resolution (3600 volumes per brain) long-duration (44 minutes) rs-fMRI scans (Fig. 1B-D). This is approximately twice the duration typically required to identify individual connectomes reliably and reproducibly from a group (42,48). The imaged volumes were parcellated into 25 regions of interest (ROI) and six functional networks representing the canonical brain networks (Fig 1B) (46,61). Notably, the largest functional connectivity differences were observed in DMN-SN, specifically between the left medial prefrontal cortex and the bilateral insula connectivity (Fig 1B-C). We also parcellated the brain into 6923 ROIs (48) and plotted the number of connections showing increased connectivity for the bipolar sibling (thresholded at 1 standard deviation across connections). This analysis revealed higher global connectivity (and decreased negative connections) in the patient brain as compared to the brain of the unaffected sibling, with the effect most consistent in the bilateral anterior insula of the salience network (Fig 1C-D). These results are consistent with the results of our previous study that negative functional resting-state brain connectivity is disrupted in DMN-SN circuitry in individuals with PBD (36). Other MRI studies on individuals with bipolar disorder observed similar connectivity deficits (62–64).

To gain insights into the cellular and molecular deficits that could be associated with disrupted brain connectivity in the PBD individual, we generated multiple iPSC lines (Supplementary Fig. 1 and Supplementary Table 2) and iPSC-derived telencephalic organoids from the recruited individuals (Fig. 2). Patient and control telencephalic organoids were produced from iPSC-derived single neural rosettes (SNR) (Fig 2A-B). We previously demonstrated that 1-month-old SNR-derived organoids consist of telencephalic neural progenitors and cortical excitatory and lateral ganglionic eminence-derived inhibitory neurons (59). To identify the differentially expressed genes (DEGs) and overrepresented pathways in patient organoids, we performed bulk RNA sequencing on twelve organoids obtained from each member of the quartet (three organoids per individual) and pair-wise comparisons using Gene Set Enrichment Analysis (GSEA) on all DEGs (Supplementary Fig. 3 and Supplementary Dataset 1) (57). We categorized the top 15 up-and down-regulated Gene Ontology (GO) gene sets according to normalized enrichment score for patient vs. unaffected sibling (Fig 2C) and patient vs. mother, father, and all controls combined (Supplemental Fig 4, and Supplementary Dataset 1). The enrichment analysis revealed that patient organoids showed significant overrepresentation of DEGs in the “intermediate filament-based process”, “positive regulation of synapse assembly”, “neuron projection extension involved in neuron projection guidance”, “regulation of axon guidance”, “inhibitory synapse assembly” GO pathways (Fig 2C and Supplemental Fig 4).

**Figure 2.**
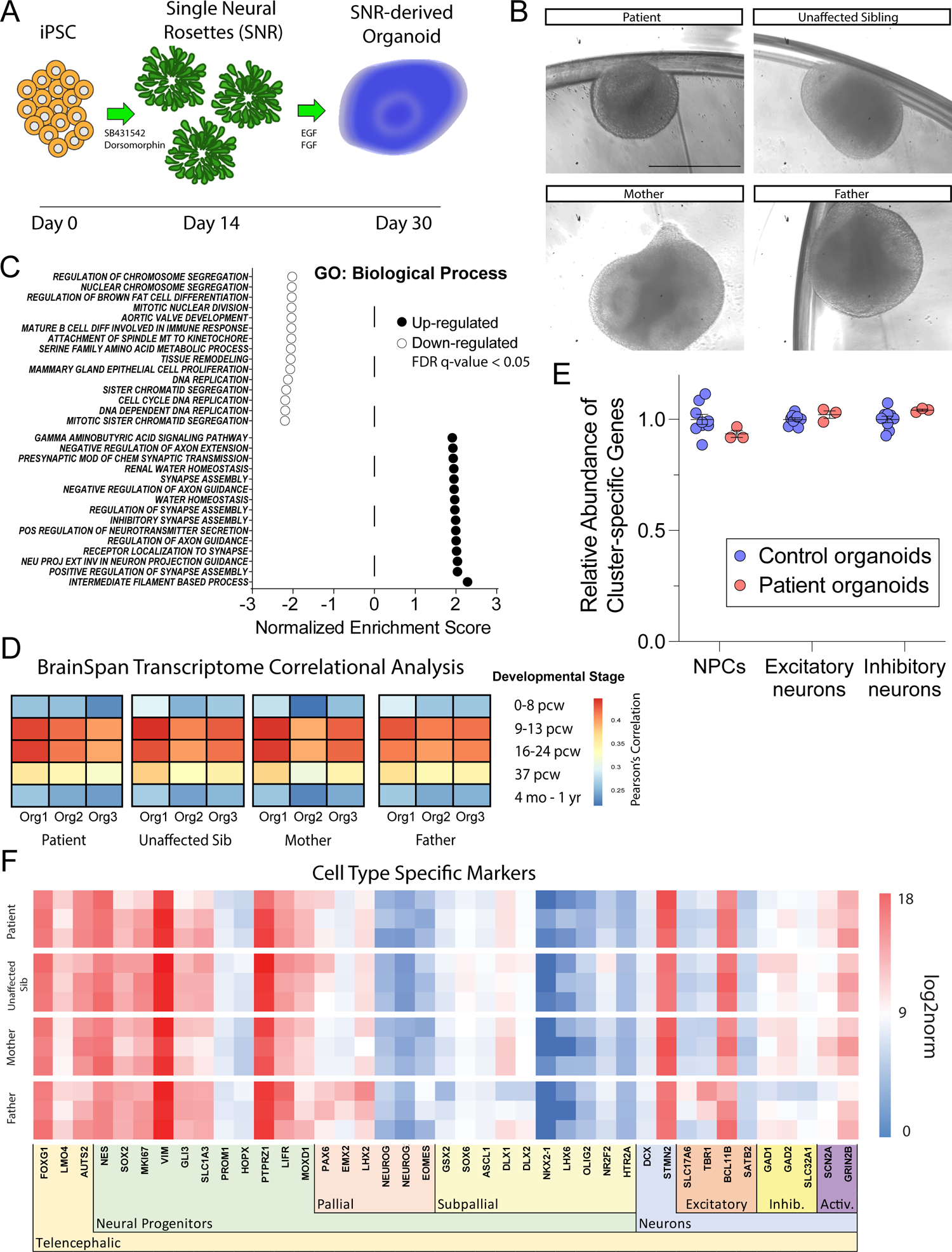
Dysregulated gene expression in gene ontology pathways related to neurite outgrowth in patient iPSC-derived organoids. A. Protocol schematic for generation of iPSC-derived organoids. B. Representative images of 1-month-old organoids used for bulk RNA sequencing. Scale bars, 500 μm. C. Gene Set Enrichment Analysis (GSEA) of top 15 most up- and down-regulated pathways for Gene Ontology gene set “GO: Biological Process” (FDR q<0.05). D. Heatmap depicting Pearson’s Correlation coefficients of patient and control organoids (3 per line) with BrainSpan transcriptome of cortical human tissue at different fetal development stages (www.brainspan.org). E. Comparison of relative cell-type composition in patient (n=3) and control organoids (n=9) using scBio bulk RNA sequencing deconvolution algorithm (58). F. Heatmap visualization of normalized gene expression of selected cell type-specific marker genes in patient and control organoids.

One possible explanation for these transcriptomic differences could be due to differences in the developmental age and/or cellular composition between patient and control organoids. To assess the developmental age of patient and control organoids, we calculated the Pearson correlation coefficient between each organoid and human cortical transcriptome at multiple developmental time points from the BrainSpan dataset (65). The gene expression profiles in both patient and control SNR-derived organoids correlated the strongest with the transcriptome of early embryonic stages 9-24pcw of the fetal brain and did not appear to differentiate patient and unaffected family organoids (Fig 2D). This suggests that patient and control organoids model early fetal developmental stages and that patient organoids do not show apparently accelerated or delayed developmental maturation as compared to the controls.

To examine the relative proportion of progenitor cells, excitatory, and inhibitory neurons, we performed Cell Population Mapping (58) of bulk RNA sequencing data based on single-cell RNA sequencing gene expression profiles obtained in our previous study on 1-month-old SNR-derived organoids (Fig. 2E). This analysis confirmed that there are no significant differences in the relative proportions of neural progenitors, excitatory neurons, or inhibitory neurons between patient and control SNR-derived organoids. We further corroborated these results by examining the expression levels of multiple cell-type-specific markers, including those that are expressed in pallial (PAX6, EMX2, LHX2, NEUROG, EOMES) and subpallial (GSX2, SOX6, ASCL1, DLX1, DLX2, NKX2.1, LHX6, OLIG2, NR2F2, HTR2A) neural progenitors (NES, SOX2, MKI67, VIM, GLI3, PROM1, HOPX, PTPRZ1, LIFR, MOXD1), as well as excitatory (SLC17A6, TBR1, BCL11B, SATB2) and inhibitory (GAD1, GAD2, SLC32A1) neurons (DCX, STMN2, SCN2A, GRIN2B) (Fig 2F). We found no apparent differences in the expression levels of these cell-type-specific markers compared to unaffected organoids. Together, these results indicate that patient and control organoids model similar early developmental stages and consist of similar types of cells, but may have deficits related to neurite development, axon guidance, or synapse assembly.

To investigate neurite development in patient and control neurons, we generated telencephalic iPSC-derived neurons from 3 patient and control iPSC lines (Fig. 3). To characterize the identities of differentiated cells from patient and control iPSCs, we performed immunostainings with antibodies against a proliferation marker Ki67 (Fig. 3A), deep layer cortical marker TBR1 (Fig. 3B), superficial layer cortical marker SATB2 (Fig. 3C), and inhibitory neuron marker GAD67 (Fig. 3D). Consistent with the bulk RNA-seq results obtained on organoids, we observed similar proportions of different cell types among control and patient iPSC-derived cells. We also observed that the majority (60-75%) of differentiated neurons were characterized by TBR1 expression, suggesting deep layer cortical identity. We next investigated the total neurite length in 1-month-old patient and control neurons that were replated at low-density, 48 hours post-replating (Fig 3E-H). Interestingly, we discovered that patient iPSC-derived neurons demonstrated significantly reduced neurites as compared to control neurons, suggesting that impaired neurite outgrowth detected in patient neurons could be the earliest cellular endophenotype associated with disrupted gene expression networks detected in patient organoids and functional brain connectivity deficits observed in the patient brain.

**Figure 3.**
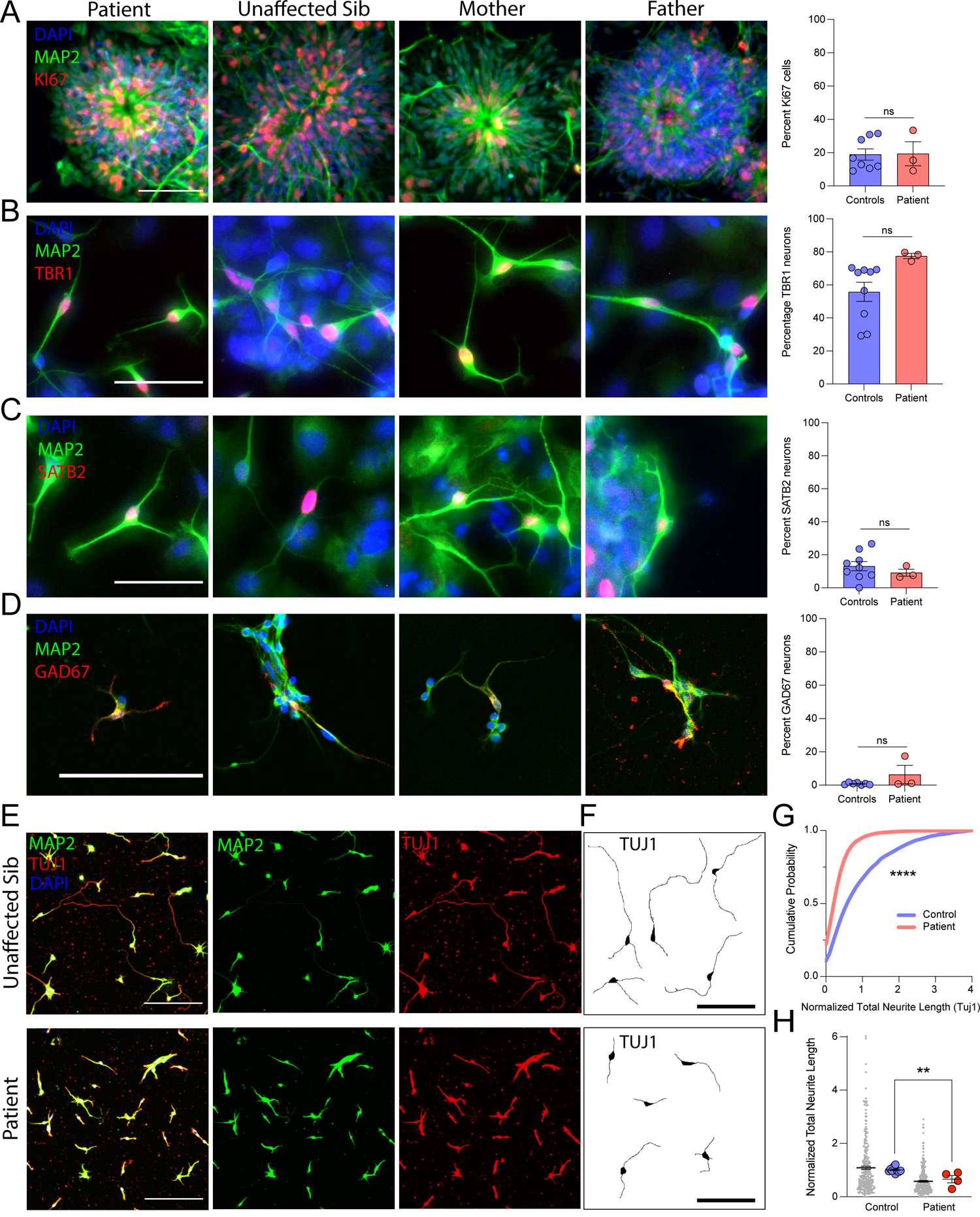
Impaired neurite outgrowth in patient iPSC-derived cortical neurons. A-D. Representative images (left) and quantifications (right) of different cell types in cultures of patient and control iPSC-derived neurons using immunostainings with cell-type-specific antibodies against KI67, MAP2, and DAPI (n=3 patient and 9 control [obtained from 3 unaffected sibling, 3 mother, and 3 father] iPSC lines in 1 differentiation/140 – 5030 DAPI positive cells per line counted) (A); TBR1, MAP2, and DAPI (n=3 patient and 9 control [obtained from 3 unaffected sibling, 3 mother, and 3 father] iPSC lines in 1 differentiation/20– 125 MAP2 expressing neurons per line counted) (B); SATB2, MAP2, and DAPI (n=3 patient and 9 control [obtained from 3 unaffected sibling, 3 mother, and 3 father] iPSC lines in 1 differentiation/48– 181 MAP2 expressing neurons per line counted) (C); GAD67, MAP2, and DAPI (n=3 patient and 9 control [obtained from 3 unaffected sibling, 3 mother, and 3 father] iPSC lines in 1 differentiation/116 – 1359 DAPI positive cells per line counted) (D). Scale bars = 100 μm. Data presented as mean + s.e.m. Unpaired t-test used to assess significance. E. Representative images of control (top) and patient (bottom) iPSC-derived neurons immunostained for MAP2, TUJ1, and DAPI. F. Representative images of traced neurons based on TUJ1 immunostaining. Scale bars = 100 μm (E-F). G. Cumulative frequency distribution of normalized total neurite length measurements in control and patient neurons (n= 6059 control [3-4 lines per individual/ 1 differentiation] and 4964 patient [4 lines/ 1 differentiation] neurons counted). ****P<0.0001 Kolmogorov-Smirnov test. H. Quantification of normalized total neurite length in patient and control neurons (small gray dots represent normalized individual total neurite measurements (n=50 randomly selected measurements per experiment for visualization), large circles represent average neurite length measurements across independent experiments. **P<0.01 Unpaired t test. All data presented as mean + s.e.m.

### Rare variant identification and prioritization

Given the highly familial expression of severe neuropsychiatric illnesses in this family and the known contributing roles of rare and ultra-rare variants in neuropsychiatric disease risk (26,66,67), we next sought to explore whether the patient may harbor rare genetic variants that may contribute to the neurite outgrowth phenotypes detected in patient iPSC-derived neurons. We performed whole-exome sequencing on the quartet and rare variant discovery and prioritization using the pVAAST/Phevor pipeline (55,56,68,69) (Fig 4A and Supplementary Dataset 2). QC analysis confirmed the quality of the sequencing data, sex and familial relationship of the sequenced quartet, and their likely European origin (Supplemental Fig 4). Using pVAAST, we identified 44 rare variants under the recessive and 0 variants under the *de novo* model of inheritance (pVAAST CLRT_p_<0.05) (Supplementary Dataset 3). We next used Phevor to re-rank the list of 44 recessive variants based on known relevance to Human Phenotype Ontology terms related to Bipolar Disorder (Behavioral abnormality HP:0000708; Abnormal emotion/affect behavior HP:0100851; Bipolar affective disorder HP:0007302) (Supplementary Dataset 3-5). We then used Polyphen (70) to identify variants that are damaging with allele frequency less than 0.05 (equivalent to 5%). This analysis yielded 15 hits, including *TRIOBP, MN1, NBPF26, PLXNB1, ANGPTL8, DOCK6, SPDYE5*, and *CEP164* (Supplementary Dataset 6).

**Figure 4.**
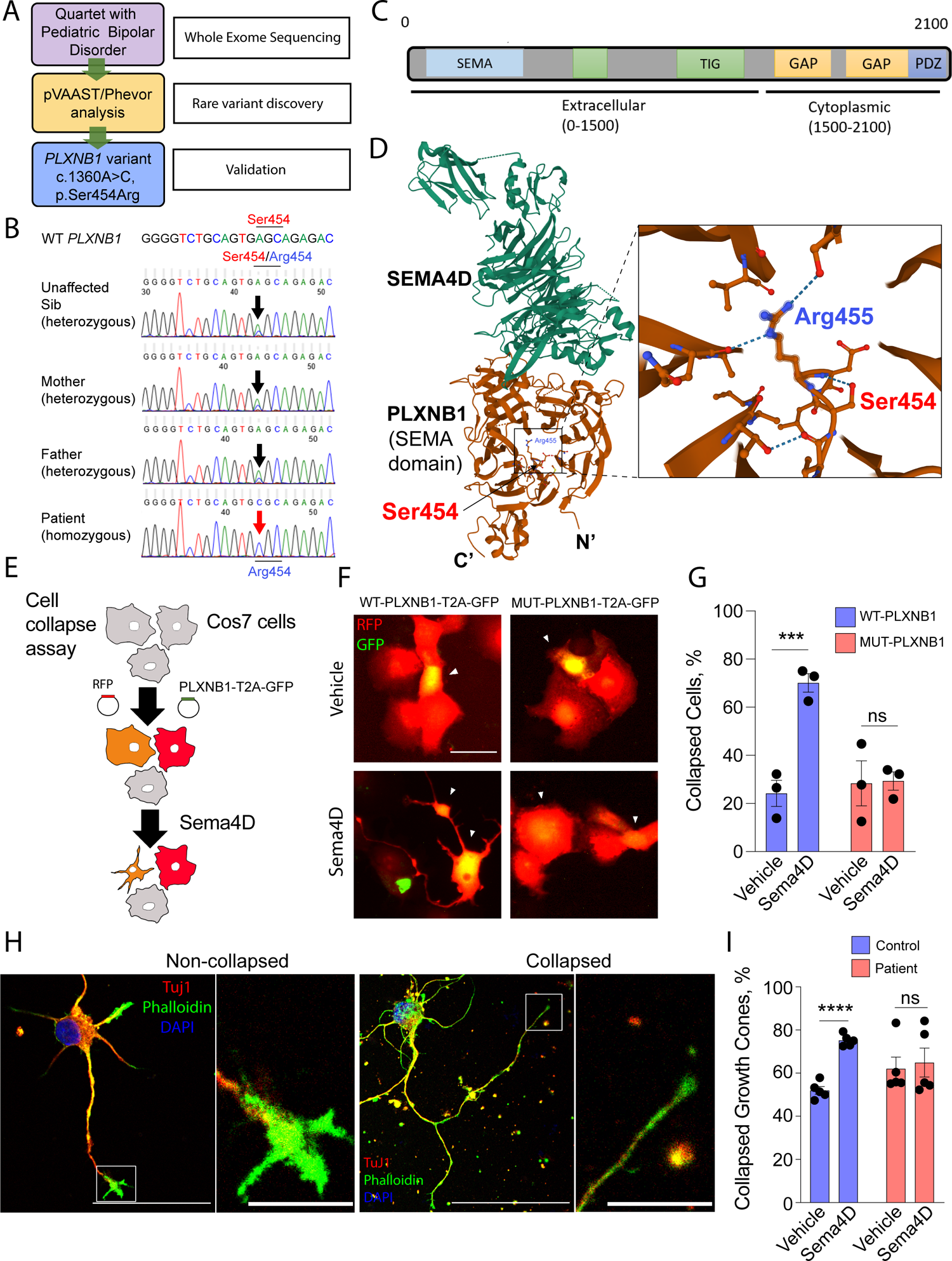
Homozygous loss-of-function mutation in PLXNB1 identified in PBD patient. A. Schematic depicting pipeline for rare variant discovery in recruited family. Whole exome sequencing was performed on proband with PBD, unaffected mother, father, and sibling. Rare variant discovery was performed using pVAAST (55), and variant prioritization was performed on the list of pVAAST hits (CLRT_p_<0.05) using Phevor (56) with HPO terms: Behavioral abnormality (HP:0000708); Abnormal emotion/affect behavior (HP:0100851); Bipolar affective disorder (HP:0007302). B. Sanger sequencing validation of homozygous PLXNB1 variant (1360A>C, pSer454Arg) in PBD patient. C. Cartoon showing PLXNB1 functional domains. PLXNB1 variant is located in the extracellular SEMA domain. D. PLXNB1-SEMA4D protein structure depicting Ser454Arg variant location. Images created using RCSB PDB viewer (https://www.rcsb.org/3d-view/3OL2) (107) for PDB ID: 3OL2. (108) E. Schematic depicting the Cos7 collapse assay. F. Representative images of Cos7 cells transfected with wild-type or mutant PLXNB1 and treated with either 2nM IgG (vehicle) or 2nM Sema4D. White arrows indicate GFP- and RFP-expressing cells. Scale bar = 100 μm. G. Quantification of the percentage of cells showing “collapsed” morphology while expressing wild-type or mutant PLXNB1, treated with vehicle or Sema4D (n=3 independent experiments/36-119 GFP- and RFP-positive cells per condition). ***P<0.001, unpaired t test. H. Representative images of neurons used for growth cone collapse analysis exhibiting “non-collapsed” (left) and “collapsed” (right) growth cone phenotypes, immunostained with TUJ1, Phalloidin, and DAPI. Scale bars = 50 μm (left), 8 μm (right). I. Quantification of percentage growth cone collapse in patient and control neurites treated with PBS (vehicle) or 8 nM Sema4D (n=5 independent experiments/3 iPSC lines from patient and unaffected sibling/10-1069 neurites per line per condition). ***P<0.001 unpaired t-test.

Of all identified variants, a homozygous variant in *PLXNB1* (3-48463799-T-G, c.1360A>C, pSer454Arg; AF=0.01322) (Fig 4B) was particularly interesting because it consistently scored in the top 6 for all Phevor rankings (Supplementary Dataset 3-5), and *PLXNB1* is an important regulator of axonal pathfinding, neurite outgrowth, and inhibitory synapse formation (71–77). In addition, a different variant in *PLXNB1* has been previously detected in a large Utah family with suicide (78), which is an extreme outcome associated with pediatric bipolar disorder (79). We also found enrichment of this *PLXNB1* variant in a large cohort of patients who died by suicide (Supplementary Dataset 7). *PLXNB1* resides in the 3p21 loci, which is highly associated with psychiatric disease risk (80), and it has two known co-receptors ErbB2 and Met (81), which were previously implicated in risk for bipolar disorder and autism in large-scale genome-wide or exome-wide association studies (24,82,83).

*PLXNB1* is a member of the plexin family of axon pathfinding proteins that act as high-affinity and high-specificity receptors for semaphorins (84,85). PlexinB1 (PLXNB1) is the receptor for Semaphorin 4D (SEMA4D) (85). Interestingly, the homozygous variant Ser454Arg identified in patient PLXNB1 resides next to positively charged residue Arg 455 in the SEMA domain (Fig 4C-D), which is critical for SEMA4D binding (85). To investigate the functional effect of this variant on PLXNB1 function, we utilized a previously well-established cell collapse assay using COS-7 cells (86,87). We transfected wild-type (WT) or mutant (MUT) PLXNB1-T2A-EGFP plasmids into COS-7 cells (Fig 4E) and investigated morphological changes in response to a transient treatment (30 mins) with 2nM SEMA4D or vehicle (Fig 4F). We observed that WT-PLXNB1-expressing cells demonstrated the expected collapsed phenotypes in approximately 70% of cells upon SEMA4D treatment (Fig 4F-G); however, no morphological changes were detected in MUT-PLXNB1-expressing cells, as compared to those treated with vehicle (Fig 4G). As we found no differences in the expression levels of *PLXNB1, PLXNB2*, or *SEMA4D* in control and patient organoids using bulk RNA sequencing (Supplementary Dataset 1), these results indicate that the Ser454Arg variant in *PLXNB1* is likely a loss-of-function mutation that impairs *PLXNB1* function mediated by SEMA4D.

To test *PLXNB1* function in patient iPSC-derived neurons, we treated both iPSC-derived neurons obtained from unaffected sibling and patient with SEMA4D (8nM for 1 hour) and investigated the proportions of collapsed growth cones (Fig. 4H-I). Consistent with the results obtained on COS-7 cells, control neurites treated with SEMA4D demonstrated a significantly increased proportion of collapsed growth cones as compared to control neurites treated with vehicle (Fig. 4I). However, no differences in the proportion of collapsed growth cones were detected in patient neurites treated with SEMA4D and vehicle (Fig 4I). Together, these results suggest that the homozygous Ser454Arg mutation in PLXNB1 is a loss-of-function mutation that may contribute to impaired neurite outgrowth in patient neurons.

### Ser454Arg mutation in PLXNB1 causes neurite outgrowth deficits

Multiple signaling cascades have been implicated in regulating neurite outgrowth and complexity in mammalian neurons (88,89). To determine whether Ser454Arg mutation in *PLXNB1* is the causative mutation for neurite outgrowth deficits detected in patient neurons, we performed rescue experiments by expressing WT-PLXNB1 in patient iPSC-derived neurons and the phenocopy experiments by expressing MUT*-* or WT-PLXNB1 in mouse cortical neurons obtained from *PLXNB1* knock out (PlxnB1-/-) mice (90) (Fig. 5).

**Figure 5.**
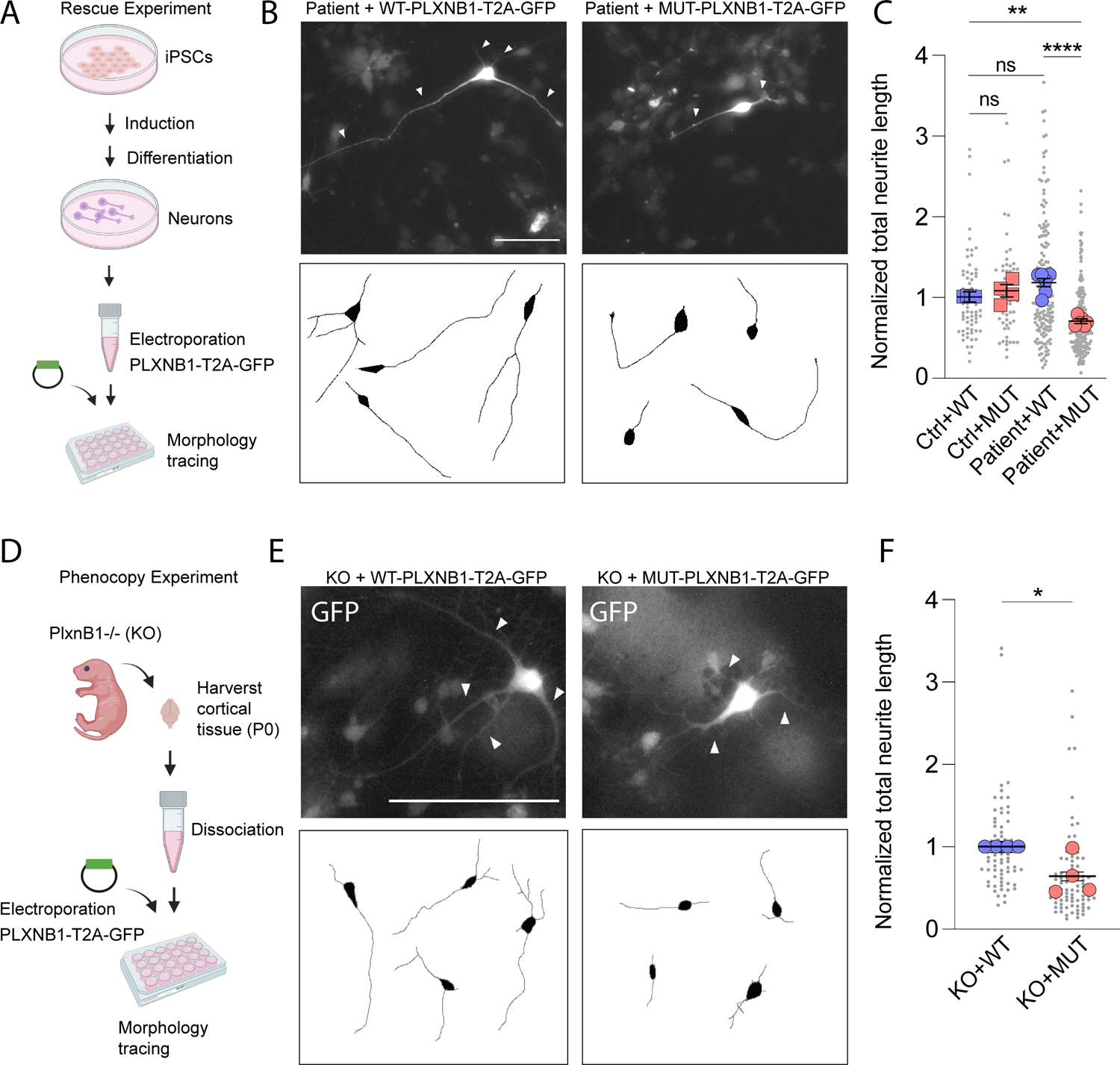
Homozygous PLXNB1 Ser454Arg mutation causes neurite outgrowth deficits. A. Schematic of PLXNB1 rescue experiment in iPSC-derived neurons. Patient and unaffected sibling (control) iPSCs underwent induction and directed differentiation. Patient and control iPSC-derived neurons were dissociated into single-cell suspension and electroporated with wild-type or mutant PLXNB1-T2A-EGFP plasmids. Neurite outgrowth was quantified 48 hours post-electroporation. B. Representative images (top) and traced (bottom) patient iPSC-derived neurons expressing GFP. White arrows indicate neurites. C. Quantification of normalized total neurite length in patient and control neurons electroporated with wild-type or mutant PLXNB1-T2A-EGFP plasmids. Small gray dots show individual neurite length measurements (n= 58-189 neurons per condition), Large shapes represent averages from independent experiments (n=3-5 independent experiments using 3 iPSC lines from patient and 3 from unaffected sibling). **P<0.01, ****P<0.0001, One-way ANOVA with Tukey’s multiple comparisons test. D. Schematic of PLXNB1 Ser454Arg phenocopy experiment. Cortical tissue from P0 PLXNB1 knock out (KO) mice was harvested and dissociated into single-cell suspension. Cells were electroporated with either wild-type or mutant PLXNB1-T2A-EGFP plasmids. Neurite length of GFP-expressing neurons was quantified 48 hours post-electroporation. E. Representative images of GFP-expressing PLXNB1-KO neurons electroporated with either wild-type or mutant PLXNB1. F. Quantification of normalized total neurite length in PLXNB1-KO neurons expressing either wild-type or mutant PLXNB1. Gray small dots represent individual total neurite measurements (n= 67 and 89 GFP-expressing neurons), Large shapes represent averages from independent experiments (n=4 independent experiments). Scale bars = 100 μm (B, E). Data presented as mean + s.e.m. *P<0.05, unpaired t-test.

As all unaffected family members recruited to this study are heterozygous for Ser454Arg in *PLXNB1*, we reasoned that the expression of WT-PLXNB1 in patient iPSC-derived neurons should rescue or compensate the neurite outgrowth deficits if it is causally related. We electroporated patient and control iPSC-derived neurons with either WT- or MUT-PLXNB1 and investigated neurite outgrowth in electroporated neurons 48 hours post-electroporation (Fig 5A). Remarkably, the total neurite length in patient neurons expressing WT-PLXNB1 was similar to the total length of neurites in control neurons expressing WT-PLXNB1 (Fig 5B-C); however, it was significantly longer than the total length of neurites in patient neurons expressing MUT-PLXNB1 (Fig 5B-C). These results indicate that Ser454Arg is a loss of function mutation and homozygous Ser454Arg mutation in *PLXNB1* is necessary for neurite outgrowth deficits detected in patient neurons.

Given that the rescue experiments were performed on human iPSC-derived neurons obtained from a single family with similar genetic background, it is conceivable that the observed neurite outgrowth deficits and the rescue are dependent on the genetic background. To test whether Ser454Arg mutation in *PLXNB1* is sufficient to cause neurite outgrowth deficits in a genetic background independent manner, we expressed WT- or MUT-PLXNB1 in cortical neurons obtained from P0 PlxnB1-/- mice (Fig. 5D). We found that the total neurite length of MUT-PLXNB1-expressing neurons is significantly shorter than that in WT-PLXNB1-expressing neurons (Fig. 5E-F). These results suggest that homozygous Ser454Arg mutation in *PLXNB1* is sufficient to cause neurite outgrowth deficits in mammalian neurons independent of the patient’s genetic background.

## Discussion

Bipolar Disorder is among the most common severe psychiatric illnesses in the world, yet the cellular and molecular deficits pertaining to this disease remain poorly understood. To gain insight into the circuit-level disruptions associated with BD, researchers have primarily taken the approach of using functional neuroimaging techniques such as fMRI. These studies have linked BD diagnoses with disrupted cortico-limbic connectivity (91,92). Postmortem studies have implicated neuroinflammation as a potential risk factor for BD(93). Genome-wide association studies have identified ∼64 significant loci associated with BD, including genes linked to ion channel and neurotransmitter receptor functions (24). Although informative on a population level, these studies have not yet produced clinically beneficial results for individual BD patients. Studies using iPSC-based models generated from BD patients have uncovered cellular deficits related to various aspects of neurodevelopment including neuronal proliferation, differentiation, cell adhesion, and neuronal excitability (15–19). However, it is unclear whether these patients also possess abnormal neural circuitry or harbor causative high-impact genetic variants.

In this study, we bridge these research designs, presenting a new combinatorial approach that leverages family-based recruitment, rs-fMRI, whole-exome sequencing, and human iPSC-derived neurons for studying the cellular and molecular deficits in patients with psychiatric illnesses. Using this approach, we identified that a rare mutation in *PLXNB1* caused neurite outgrowth deficits in iPSC-derived neurons from a patient with PBD and disrupted brain connectivity. We believe that this approach holds tremendous potential for identifying causal rare variants in PBD and other psychiatric disorders and elucidating the cellular and molecular deficits.

This is the first study to use an iPSC-based approach to model PBD, and the first study to observe neurite outgrowth phenotypes in a patient with PBD. The association between impaired neurite outgrowth and bipolar disorder is not surprising, given that bipolar disorder treatments, such as lithium, valproate, lamotrigine, and quetiapine, are known to target pathways that regulate neurite development and axonogenesis (94,95). In addition, neurite outgrowth deficits were previously implicated in BD and other neuropsychiatric disorders, including schizophrenia (96), autism spectrum disorder (97), and ADHD (98). Thus, impaired neurite outgrowth may be an early cellular phenotype that could contribute to nonspecific risk for psychiatric disease. Interestingly, most of the neurons generated from 2D culture were identified as TBR1-expressing deep-layer cortical projection neurons, which are known to establish connections with subcortical structures (99). It is conceivable that impaired neurite outgrowth in the deep-layer cortical projection neurons may disrupt brain connectivity by receiving and providing fewer input and output projections (100). However, this finding does not necessarily suggest that impaired neurite outgrowth directly causes bipolar disorder or disrupted brain circuitry in this patient. As neurite outgrowth deficits have been implicated in other neuropsychiatric disorders, this finding supports the hypothesis that neurite outgrowth abnormalities may broadly be associated with underlying risk for neuropsychiatric pathologies. Although it is unknown whether this phenotype is present in the brain, the robust observation of these deficits *in vitro* could represent an early endophenotype for drug screening for the development of novel therapies potentially important across multiple psychiatric diagnoses.

To our knowledge, this is also the first study to identify a rare genetic variant in *PLXNB1* in a patient with PBD. Interestingly, the *PLXNB1* co-receptor ErbB2 was previously identified as a BD risk gene (23), indicating a possible involvement of PLXNB1-ErbB2 signaling in BD risk. We demonstrated that *PLXNB1* Ser454Arg is a loss-of-function variant and that the homozygous *PLXNB1* Ser454Arg variant causes neurite outgrowth deficits. This finding is surprising, given that Sema4D-PlexinB1 signaling is known to cause acute growth cone collapse and reduced neurite outgrowth in mouse neurons. (101,102) However, prolonged Sema4D stimulation of PlexinB1 receptors can also induce secondary neurite branching and outgrowth. (103) Thus, the exact effects of PlexinB1 on neurite outgrowth are complex and context dependent. Although the molecular mechanisms that are responsible for this neurite outgrowth phenotype remain unknown, one hypothesis is that *PLXNB1* Ser454Arg impairs Sema4D-PlexinB1 binding and affects the *PLXNB1*-mediated downstream signaling cascades, including R-Ras and Rho-dependent pathways (72,104). Additionally, it is conceivable that the Ser454Arg mutation, which resides in the sema domain, impairs PlexinB1 binding affinity and specificity to Sema4D, causing inappropriate engagement of other ligands or constitutive activation. Intriguingly, deletion of the extracellular region of PlexinB1 causes impaired neurite outgrowth through an R-Ras-dependent mechanism (72). Furthermore, R-Ras signaling inhibits the PI3K/Akt/GSK3β/CRMP2 pathway, which is a target of lithium, and abnormal CRMP2 was previously identified in post-mortem tissue and iPSC-derived neurons of BD patients. (95) Intriguingly, lithium treatment also disrupts semaphorin signaling. (95) Taken together, this suggests that GSK3β/CRMP2 may be a convergent biological pathway affected in BD. Indeed, the high-confidence BD risk gene *ANK3* also regulates neuronal microtubule dynamics through CRMP2. (105) Future studies will be required to elucidate how the *PLXNB1* Ser454Arg variant caused neurite outgrowth deficits and whether downstream GSK3β/CRMP2 is affected. Further, although *PLXNB1* is known to regulate growth cone and neurite morphology *in vitro* (72,102), mutant mice lacking PlexinB1 are relatively normal with no major cytoarchitectural abnormalities or migration deficits (106), suggesting that major defects in PlexinB1 function *in vitro* likely result in subtle or hard-to-detect *in vivo* phenotypes. Furthermore, *PLXNB1* is known to regulate GABAergic synapse formation (73,74). It is unclear whether *PLXNB1-*related neurite outgrowth deficit or *PLXNB1-*mediated synaptic deficits contribute to disrupted connectivity in bipolar disorder. Additional studies will be required to clarify these questions.

In summary, we present in this study a new paradigm to gain mechanistic insight into patients with psychiatric illnesses, leveraging functional magnetic resonance imaging, genetic sequencing, and rare variant identification, RNA sequencing, and iPSC-based approaches. We demonstrate the utility of this approach in a quartet with a patient diagnosed with pediatric bipolar disorder and discovered a rare mutation in *PLXNB1* that impaired neurite outgrowth. One limitation of this approach, however, is the generalizability of findings to the larger patient population—a limitation that is inherent to studies with small numbers of patients. Thus, future studies using this approach should recruit larger families comprising multiple patients and unaffected relatives. We believe that this approach holds tremendous potential for helping discover and elucidate the relationship between novel rare variants and their biological function, and generate hypotheses as to how this can lead to disrupted molecular/cellular pathways and affect brain connectivity.

## Data Availability

All data produced in the present study are available upon reasonable request to the authors

## Acknowledgments

The authors are thankful to the family that participated in our study; to Jubel Morgan for collection of biological samples, Dr. Roland Friedel and Suzanne Paradis for sharing PLXNB1-null mice; to Dr. Xuewu Zhang for sharing PlexinB1 plasmid; to Drs. Megan Williams, Sungjin Park, Richard Dorsky, and Jan Christian for advice on experiments and comments on the manuscript; to Dr. Brian Dalley, Opal Allen, and Brian Lohman and Utah high-throughput genomics facility for help with sequencing and data analysis; to Utah cell imaging facility for help with imaging; to Dr. Elliott Farris for help with analysis of variant data; to Dr. Marvin B. Moore and the Center for high performance computing for help with processing and analysis of whole-exome sequencing data. The authors would also like to thank the funding sources that made this study possible: the Utah Neuroscience Initiative, the Utah Genome Project, and the NIH Developmental Biology Training Grant (Award Number: T32HD007491).

