## Supplementary Materials for "Neurite outgrowth deficits caused by rare PLXNB1 mutation in pediatric bipolar disorder"

**This PDF file includes:**

Supplementary Figures 1-5

Supplementary Table 1-2

Captions for Supplementary Dataset 1-7

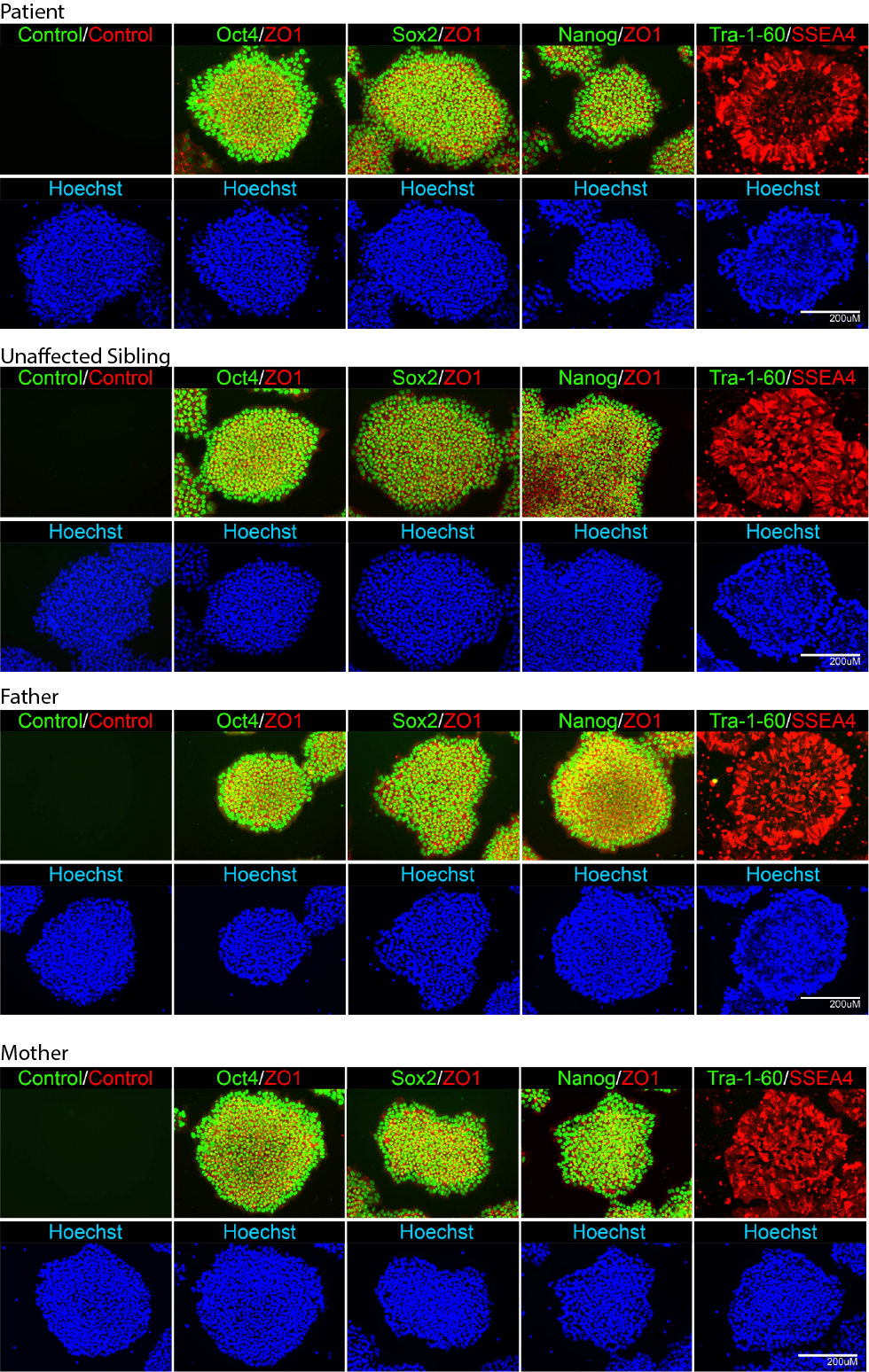

**Supplementary Figure 1. iPSC Generation and Characterization.** Representative images of iPSCs generated from patient, unaffected sibling, mother, and father immunostained with Oct4/ZO1, Sox2/ZO1, Nanog/ZO1, and Tra-1-60/SSEA4 are shown. Scale bar = 200uM.

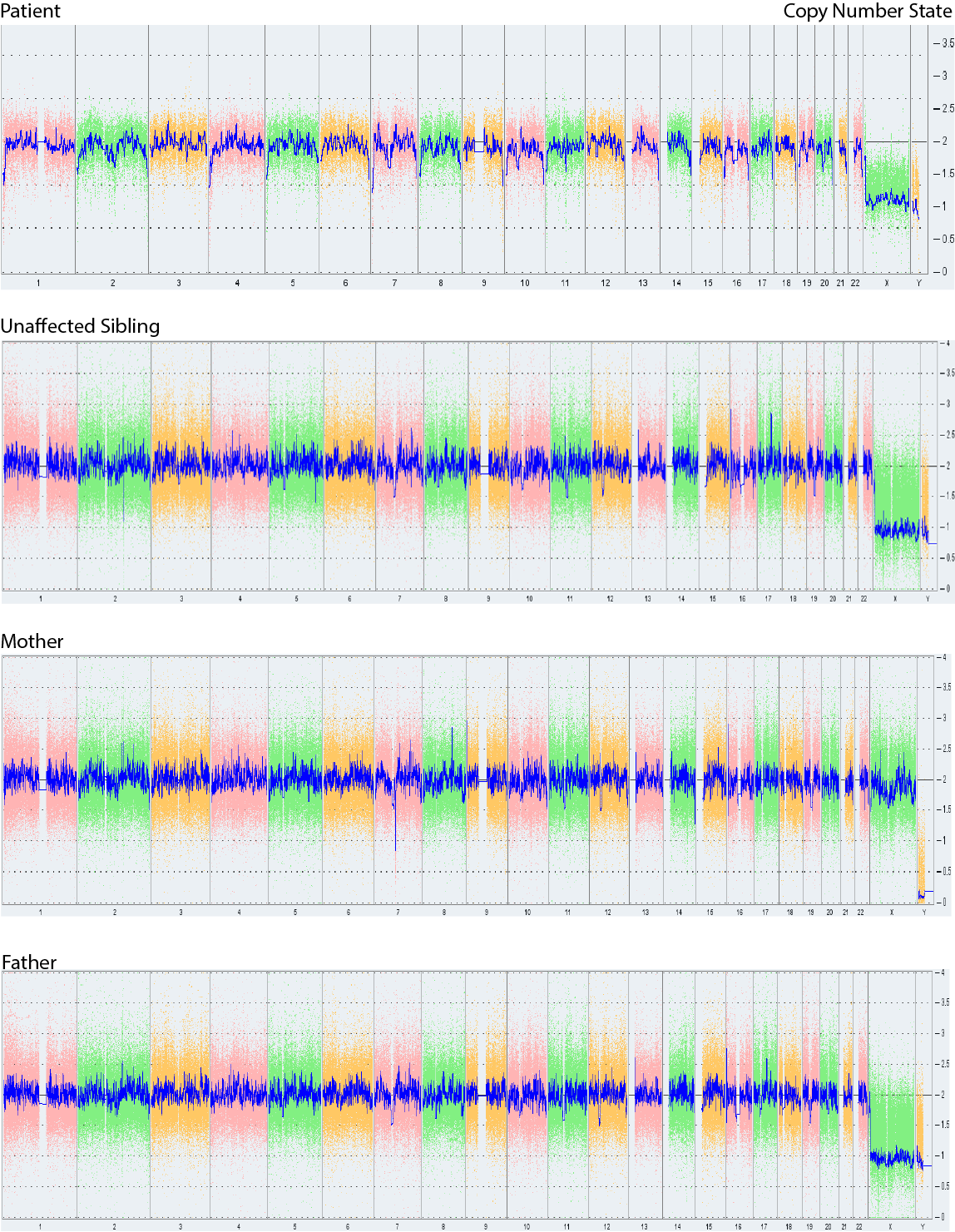

**Supplementary Figure 2. iPSC Karyotyping.** The whole genome view for patient, unaffected sibling, mother, and father iPSCs copy number status are shown. The smooth signal plot (right y-axis) is the smoothing of the log2 ratios which depict the signal intensities of probes on the microarray. A value of 2 represents a normal copy number state (CN = 2). A value of 3 represents chromosomal gain (CN = 3). A value of 1 represents a chromosomal loss (CN = 1). The pink, green and yellow colors indicate the raw signal for each individual chromosome probe, while the blue signal represents the normalized probe signal which is used to identify copy number and aberrations (if any).

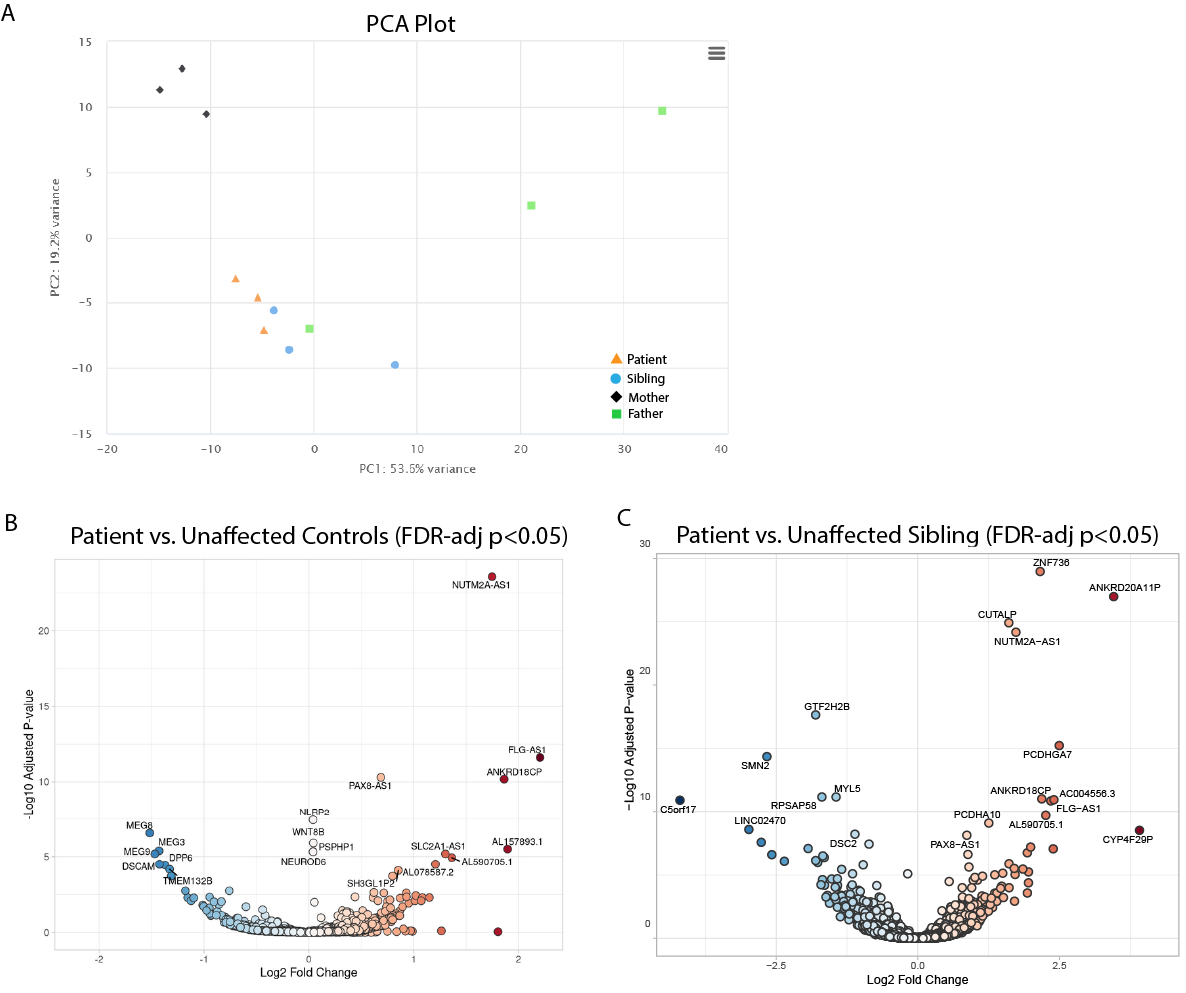

**Supplementary Figure 3. Bulk RNAseq PCA and volcano plots for patient vs. controls.** A. PCA plot for all organoids from patient (n=3) and unaffected controls (n=3 each from sibling, mother, and father). B. Volcano plot depicting DEGs from patient vs. all unaffected control organoids (FDR-adjusted p<0.05). C. Volcano plot depicting DEGs from patient vs. unaffected sibling organoids (FDR-adjusted p<0.05).

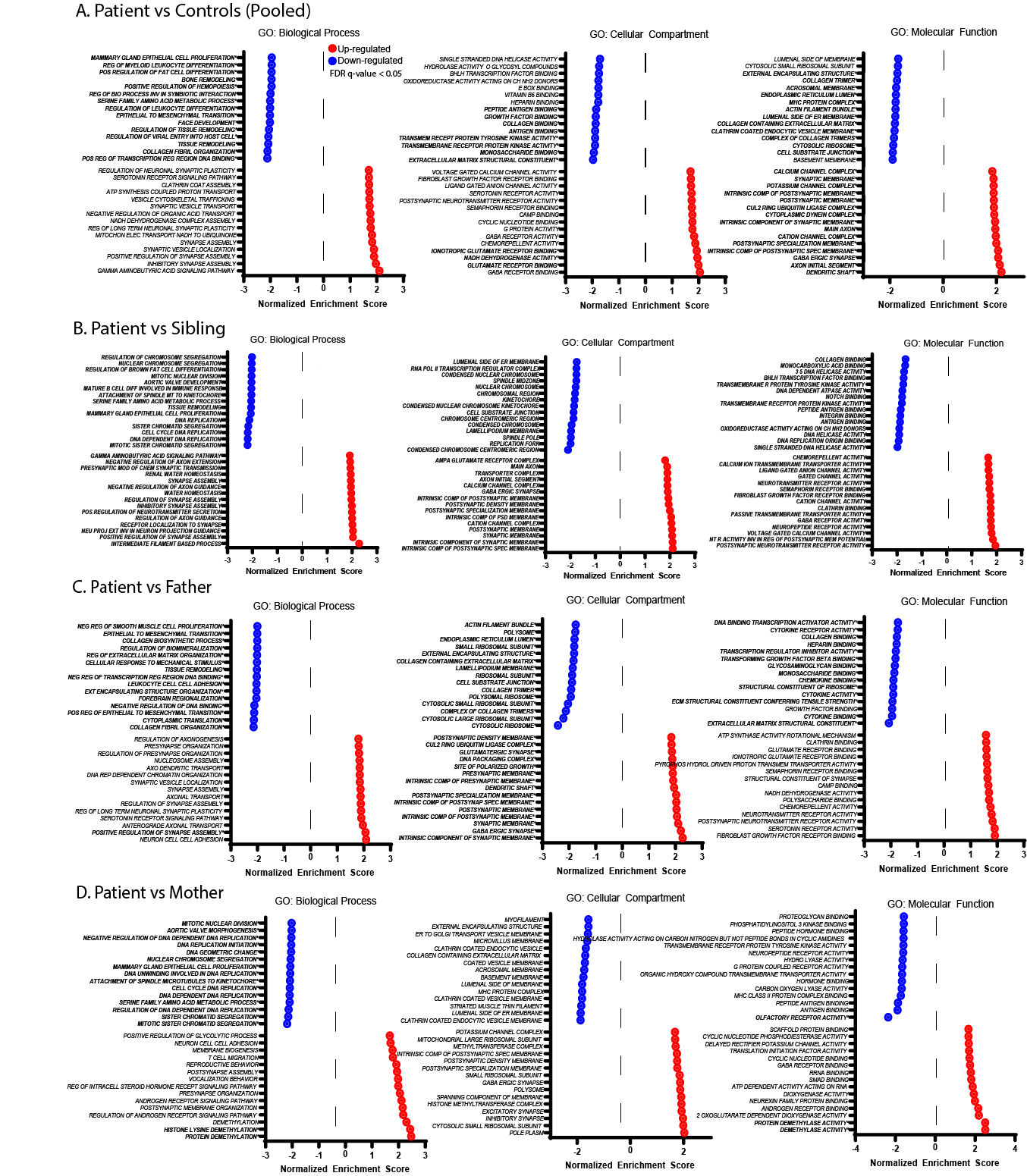
**Supplementary Figure 4. Gene Set Enrichment Analysis results for patient vs. control organoid.** A-D. Figure depicting the top 15 up- and down-regulated pathways from Gene Set Enrichment Analysis (GSEA) for patient (n=3) vs. control organoids (n=3 for each unaffected individual) using the following gene sets: GO: Biological Process, GO: Cellular Component, GO: Molecular Function. All significant gene sets (FDR-adjusted p<0.05) are depicted in bold.

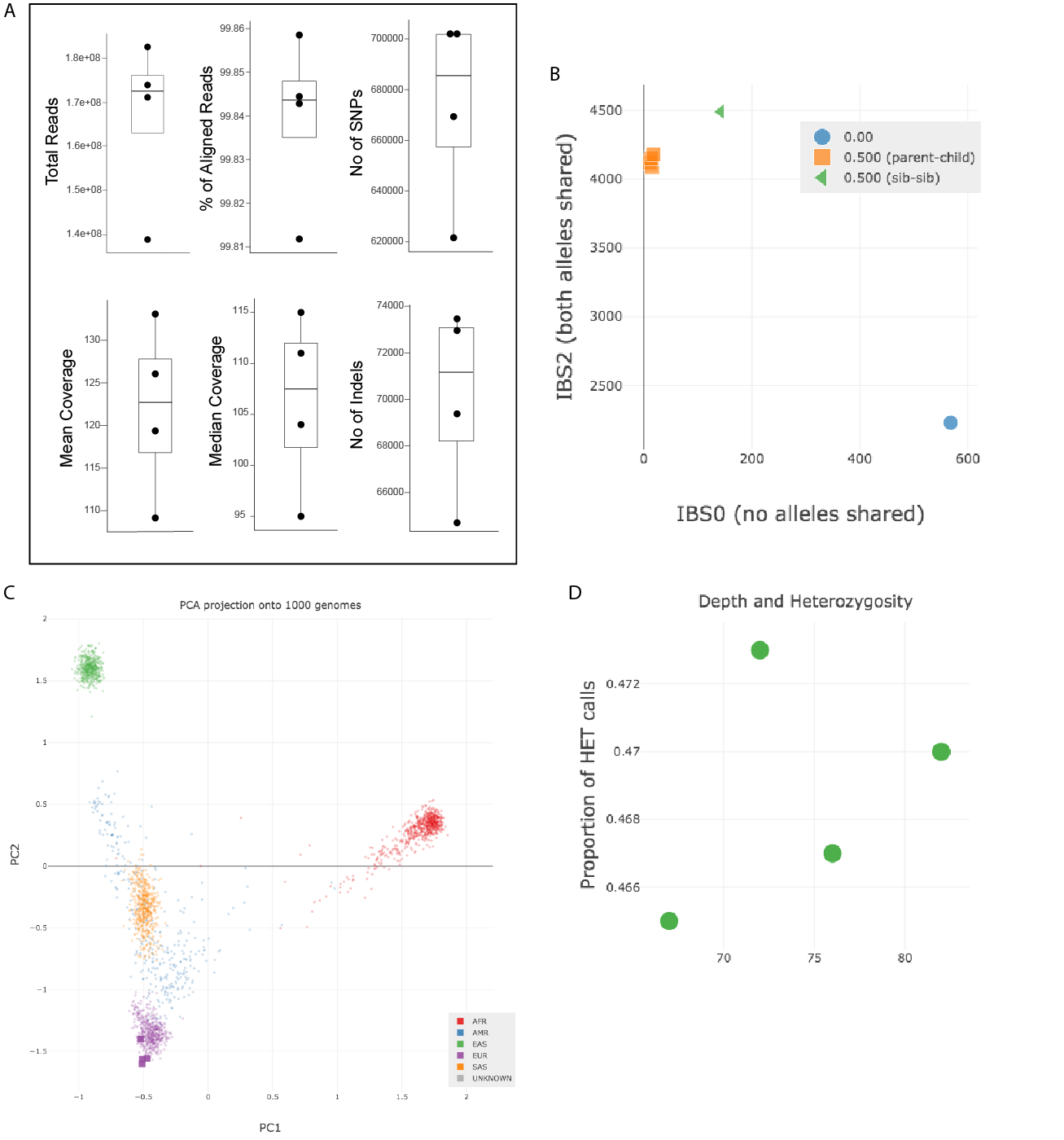
**Supplementary Figure 5. Whole Exome Sequencing QC.** A. Whole exome sequencing produced a mean of 166 million reads per individual (range 138-182 million) and an average of 99.8% mapped/aligned reads to the GRCh38 human reference genome with a duplication range between 11-20% amongst all four samples. Captured Mean and median coverage were within the expected range for WES data. Variant calling produced an average of 673,764 SNVs and 70,125 indels per sample. Peddy was used to confirm the relatedness (B), ancestry (C), and (D) and to evaluate the proportion of heterozygous calls per sample. No sample outliers were detected.

Supplementary Table 1. Assessment of WASI and CBCL in patient and unaffected sibling.

| WASI IQ Scores | | | | |
| --- | --- | --- | --- | --- |
|  |  | Sum of Scores | IQ | Percentile |
| Patient | Full-2 | 125 | 122 | 93 |
| Unaffected Sibling | Full-2 | 127 | 124 | 95 |
| CBCL Internal/External Score | | | | |
|  |  | Internalizing Problems | Externalizing Problems | Total Problems |
| Patient | Raw Score (Sum) | 19 | 6 | 41 |
|  | T score | 69-C | 51 | 60-B |
|  | Percentile | 97 | 54 | 84 |
| Unaffected Sibling | Raw Score (Sum) | 12.5 | 11.5 | 6 |
|  | T score | 55 | 59 | 55 |
|  | Percentile | 69 | 81 | 69 |

**Supplementary Table 2. Summary of iPSC lines used for the experiments detailed in this manuscript.**

|  | Patient | | | | | Unaffected Sibling | | | | | Mother | | | | Father | | | |
| --- | --- | --- | --- | --- | --- | --- | --- | --- | --- | --- | --- | --- | --- | --- | --- | --- | --- | --- |
| Clone # | 1 | 2 | 3 | 4 | 5 | 1 | 2 | 3 | 4 | 5 | 1 | 2 | 3 | 4 | 1 | 2 | 3 | 4 |
| iPSC Characterization |  | x |  |  |  |  | x |  |  |  |  | x |  |  |  |  |  |  |
| iPSC | x | x | x | x | x | x | x | x | x | x | x | x | x | x | x | x | x | x |
| NPC | x | x | x | x | x | x | x | x | x | x | x | x | x | x | x | x | x | x |
| Neuron | x | x | x | x | x | x | x | x | x | x | x | x | x | x | x | x | x | x |
| IHC (Satb2/Tbr1) |  | x |  | x | x |  | x |  | x | x | x |  | x | x | x |  | x | x |
| IHC (GAD67) |  | x | x | x |  |  | x | x | x |  | x | x | x |  | x | x | x |  |
| IHC (Ki67) |  | x | x | x |  |  | x | x | x |  | x | x | x |  | x | x | x |  |
| Neurite Outgrowth | x | x |  | x | x | x | x |  | x | x | x |  |  |  | x |  |  |  |
| Sema4D Growth Cone assays |  | x |  | x | x |  | x |  | x | x |  |  |  |  |  |  |  |  |
| Generation of organoids |  |  | x |  |  |  |  | x |  |  |  | x |  |  |  | x |  |  |
| Bulk RNAseq of organoids |  |  | x |  |  |  |  | x |  |  |  | x |  |  |  | x |  |  |
| PLXNB1 Rescue experiment |  | x |  | x | x |  | x |  | x | x |  |  |  |  |  |  |  |  |

Supplementary Dataset 1 (separate file). Gene Set Enrichment Analysis of patient and control organoids bulk RNA-sequencing data.

Supplementary Dataset 2 (separate file). Pathogenic variants identified through pVAAST (recessive run)

Supplementary Dataset 3 (separate file). Phevor prioritization of pVAAST recessive hits for Human Phenotype Ontology: Behavioral Abnormality (HP: 0000708). Top 30 hits are listed.

Supplementary Dataset 4 (separate file). Phevor prioritization of pVAAST recessive hits for Human Phenotype Ontology: Abnormal emotion/affect behavior (HP:0100851). Top 30 hits are listed.

Supplementary Dataset 5 (separate file). Phevor prioritization of pVAAST recessive hits for Human Phenotype Ontology: Bipolar affective disorder (HP: 0007302). Top 30 hits are listed.

Supplementary Dataset 6 (separate file). List of pVAAST variants predicted to be damaging and/or deleterious (PolyPhen). Red asterisk denotes compound heterozygotes.

Supplementary Dataset 7 (separate file). Enrichment analysis of pVAAST hits in Utah suicide database.
